# An assessment of rates and covariates of mpox diagnosis and vaccination provides evidence to refine eligibility criteria for mpox vaccination among gay, bisexual and other men who have sex with men in the Netherlands

**DOI:** 10.1101/2023.02.28.23286578

**Authors:** Philippe C.G. Adam, Eline L.M. Op de Coul, Paul Zantkuijl, Maria Xiridou, Hanna Bos, Cor Blom, Itsada Ketsuwan, Margreet J.M. te Wierik, Silke David, John B.F. de Wit

**Author notes:** **Correspondence**: John B.F. de Wit,.

## Abstract

**Background:** The 2022 multicountry mpox outbreaks predominantly affected gay, bisexual and other men who have sex with men (GBMSM) in non-endemic countries, including the Netherlands. We conducted a survey-based assessment of the alignment between the risk factors associated with mpox diagnosis among GBMSM in the Netherlands and the eligibility criteria used in 2022 for vaccinating this group, with the aim to refine these criteria.

**Methods:** An online self-report survey was conducted among adult GBMSM in the Netherlands between 29 July and 30 August 30, 2022, corresponding to the first month of the Dutch mpox vaccination campaign. GBMSM were recruited via advertisements on social media and gay dating apps. Participants reported on their sexual behaviour, mpox diagnosis, and/or (initial) mpox vaccination since the start of the outbreak. Covariables of mpox diagnosis and vaccination were assessed using logistic regression analyses.

**Results:** Of the 2,460 participants, 73 (3.0%, 95% CI 2.3%-3.6%) were diagnosed with mpox and 485 (19.7%, 95% CI 18.1%-21.3%) had received (initial) mpox vaccination. Using sample weighting, we estimated that, of the GBMSM population aged 18-80 years in the Netherlands, 1.1% (95% CI 0.7%-1.6%) had been diagnosed with mpox and 7.8% (95% CI 6.8%-8.9%) had received (initial) vaccination. HIV-PrEP use, living with HIV, reporting 20 or more sex partners in the past 12 months, and sex in sex venues/parties in the past 2 months were independent risk factors for mpox diagnosis. Except for sex in sex venues/parties, these variables were also independently associated with mpox vaccination.

**Conclusion:** This study provides novel evidence regarding the degree to which eligibility criteria for mpox vaccination align with the risk factors for mpox among GBMSM in the Netherlands. The findings contribute to a refinement of the eligibility criteria for mpox vaccination, to which sex in sex venues/parties should be added.

## Introduction

Mpox (formerly named monkeypox [1]), is a zoonotic infection caused by the monkeypox virus (MPXV) [2], that is endemic in parts of West and Central Africa [3]. With an estimated global total of about 30,000 cases in humans until 2019, its occurrence was considered rare until early 2022 [4]. Following initial reports of unusual cases in the United Kingdom in early May 2022 [5], multicountry outbreaks of human-to-human transmission of mpox have been identified in other non-endemic regions, including in Europe [6,7]. Between 1 January 2022 and 18 February 2023, 86,019 laboratory confirmed cases of mpox from 110 countries were reported to the World Health Organization (WHO) [8]. The 2022 mpox outbreaks predominantly occurred in gay, bisexual and other men who have sex with men (GBMSM) and likely resulted from transmission during sexual contact, making individuals with a higher number of sex partners and/or participating in specific sexual networks more susceptible to mpox [9,10, 11].

Observational studies suggest that smallpox vaccination attenuates mpox disease severity and acquisition risk [2,3,12], and WHO recommends primary prevention vaccination of individuals at high risk, notably GBMSM with multiple sexual partners [13]. However, as in other countries [14], the vaccine for mpox prevention is part of the strategic stockpile and was previously not available to the public in the Netherlands. This limited supply of vaccine necessitates the prioritisation of vaccination to population groups at highest risk [15]. The optimal allocation of vaccine requires an alignment of eligibility criteria for vaccination with risk factors for mpox, knowledge of which was limited when mpox vaccination programs were initiated. Based on data from the Netherlands until the end of August 2022, this study provides new evidence on risk factors related to mpox diagnosis among GBMSM and assesses to what extent eligibility criteria for mpox vaccination are aligned with risk factors for mpox diagnosis.

The Netherlands is one of the European countries most affected by the mpox outbreak [7]. As of 16 February 2023, there were 1,261 confirmed cases of mpox in the Netherlands [16], a country with nearly 18 million inhabitants [17]. The first mpox case in the Netherlands was reported on 20 May 2022, and the number of new diagnoses peaked at the beginning of July 2022. After that time, the number of new diagnoses decreased and became sporadic as of mid-September 2022 [16]. In the second half of 2022, the number of mpox cases declined in the Netherlands [16], as well as in Europe [18] and globally [8]. Future trends in mpox infections remain uncertain, although new outbreaks are likely to be of smaller magnitude than the 2022 outbreak [19,20]. In this context, mpox remains a public health concern, and achieving and sustaining elimination are priorities [21].

The early mpox response in the Netherlands aimed at halting transmission and consisted of identifying infected people and implementing public health protection measures, including isolation, contact tracing and post-exposure prophylaxis (PEP) vaccination [11]. Awareness campaigns were also conducted among GBMSM. The stepwise roll-out of a centralised pre-exposure prophylaxis mpox vaccination program, using 32,000 doses of Imvanex® third generation smallpox vaccine, was initiated on 25 July 2022 [22]. The vaccination program made use of initial information on the characteristics of individuals diagnosed with mpox [11] to prioritise which GBMSM would (first) receive mpox vaccination.

General eligibility criteria for mpox vaccination were defined during meetings of the mpox expert council [23] and further specified for practical program implementation from available data on risk indicators routinely recorded in PrEP, HIV and STI programs. This resulted in practical eligibility criteria for mpox vaccination for GBMSM and trangender persons encompassing: 1) prescribed HIV-PrEP at a sexual health centre (SHC) or by a general practitioner (GP), or registered on a waiting list for HIV-PrEP at a SHC; 2) living with HIV and receiving regular HCV screening as a proxy for high risk behaviour; or 3) having attended a SHC in the past six months because of 3a) partner notification related to HIV or STI, 3b) prior diagnosis of syphilis, gonorrhoea or chlamydia, or 3c) having had more than three partners in the past six months [22].

High-risk individuals eligible for vaccination were identified by the National Institute for Public Health and the Environment (RIVM) from the national surveillance database (SOAP) that includes information pertaining to all visits to SHCs, encompassing anonymised data on age, sex, (sexual) behaviour, STI diagnoses, HIV status, and use of HIV-PrEP (M. Haverkate, personal communication, 6 July 2023). From these data, lists of individuals eligible for vaccination were provided to each SHC by region. For persons receiving HIV-PrEP from their GP, GPs were contacted by the Public Health Services (PHS) in their region to invite eligible patients for vaccination. Similarly, the HIV healthcare providers of people living with HIV were asked by the PHS to invite eligible patients for vaccination. All eligible individuals identified received an invitation to vaccinate against mpox.

To gather comprehensive information on vaccination among individuals most at risk for mpox, we conducted an online survey among adult GBMSM in the Netherlands. Drawing on the survey data, we examined the proportions of GBMSM reporting mpox diagnoses and mpox vaccination, and the associations of mpox diagnosis and mpox vaccination with a range of potential risk factors. These factors encompassed the use of HIV-PrEP, living with HIV, prior STI diagnosis, number of sex partners, and engagement in group sex, chemsex, and sex in gay saunas, sex clubs or at sex parties. We compared the factors associated with mpox diagnosis and mpox vaccination to evaluate the degree of alignment between the risk factors for mpox, the eligibility criteria for for mpox vaccination, and the factors associated with mpox vaccination in various groups of GBMSM. Furthermore, we estimated the proportion of GBMSM reporting one or more risk factors for mpox, and estimated the level of immunity towards mpox achieved within this group due to mpox diagnosis, mpox vaccination or prior smallpox vaccination. In addition to providing novel evidence on the risk factors for mpox among GBMSM, the study findings contribute to guiding the targeting the mpox vaccination program in the Netherlands to GBMSM most at risk.

## Materials and Methods

### Design and procedures

A new, purposive cross-sectional self-report survey entitled *Monkeypox: a new challenge for your sex life* was conducted online among GBMSM in the Netherlands between 29 July and 30 August 2022. Participants were recruited via ads appropriate for the GBMSM population of interest on social media (i.e., Facebook and Instagram), gay dating sites and apps (i.e., Grindr and Recon), and *Man tot Man* the principal sexual health promotion platform for GBMSM in the Netherlands (https://www.mantotman.nl/en). The ads provided a link to a web page with information about the study. People were eligible to participate if they: lived in the Netherlands, were 18 years or older, identified as male (or non-binary or transgender) and ever had sex with a man. All participants provided informed consent and received no compensation. Ethics approval for this study was granted on 25 July 2022 by the Faculty Research Ethics Board of the Faculty of Social and Behavioural Sciences, Utrecht University, the Netherlands (reference number: 22-0358).

### Measures

*Participant characteristics:* age (continuous in years and in five age ranges: 18–29 years, 30– 39 years, 40–49 years, 50–59 years, and 60 or more years), education (tertiary education completed or ongoing, no/yes), province of residence (recoded as the Randstad metropolitan area, no/yes, that is, the main urban area of Netherlands encompassing the four largest cities, their suburbs and towns in between), sexual orientation (gay, bisexual, heterosexual, unsure). *Sexual behaviours:* number of male sex partners in the past 12 months (0, 1–9, 10–19, 20–49, 50–99, 100 or more; recoded as 0–9, 10–19, 20 or more), group sex (sex with two or more partners at the same time) in the past two months (no/yes), chemsex (the intentional use of drugs to enhance sex) in the past two months (no/yes), sex in a gay sauna, sex club or at a sex party (for brevity also referred to as sex venues or parties) in the past two months (no/yes). *HIV/STI-related indicators:* current HIV-PrEP use (no/yes), living with HIV (no/yes), STI diagnosis in the past 12 months (no/yes).

*Mpox-related indicators:* mpox diagnosis (no/yes), invitation received to vaccinate against mpox (no/yes), and having received (initial) mpox vaccination since the start of the 2022 outbreak (no/yes). Participants were not asked about the specific number of vaccine doses they had received. However, it is likely that the participants reporting mpox vaccination in this survey had only received one dose of the mpox vaccine. This is because the survey was conducted during the first month of the mpox vaccination campaign and the recommended interval between the first and second doses is at least four weeks [16].

## Data analysis

Only eligible participants who fully completed the questionnaire were retained in the analyses presented in this paper. All analyses were conducted using SPSS (version 28). Descriptive statistics were computed to describe the characteristics of the sample. Due to a lack of data regarding the characteristics of the GBMSM population in the Netherlands, it was not possible to evaluate the sample’s representativeness with confidence. Nonetheless, it was clear from a comparison with national records [24] that the sample overrepresented GBMSM who use HIV-PrEP or live with HIV. A proportional weighting procedure was therefore applied to the data to redress the overrepresentation of GBMSM using HIV-PrEP or living with HIV. Sample weights were calculated based on data from national records showing that 11,576 GBMSM nationally used HIV-PrEP, and 13,289 GBMSM were in HIV care in 2022 [24]. The size of the sexually active GBMSM population aged 18-80 years living in the Netherlands was estimated at 310,000, based on an update of a previous estimate [25]. Our update was guided by social science research data regarding same-sex attraction and behaviour as well as gay and bisexual identity in the Netherlands [26]. We also accounted for the growth of the general population of the Netherlands [27]. Details of the study population size estimation are described in the Supplemental materials.

The proportional weighting procedure kept the sample size constant compared to the unweighted sample. Per convention, numbers are only reported for the unweighted data. To assess the impact of the weighting procedure on the sample characteristics, we calculated differences in percentages between unweighted and weighted estimates and computed the ratio of the two estimates.

Descriptive statistics were also calculated to estimate the proportions of participants (and 95% confidence intervals [CI]) reporting mpox diagnosis or mpox vaccination using both unweighted and weighted data. Univariable and multivariable logistic regression analyses were used to identify the covariables associated with mpox diagnosis or mpox vaccination. The selected set of covariables included in the models encompassed both the general and specific eligibility criteria for vaccination used in the Netherlands (see Table 1). Additional sexual behaviours were also included that may be related to the transmission of MPXV but were not listed as eligibility criteria for mpox vaccination in the Netherlands in 2022. In total, seven potential covariables were assessed: current use of HIV-PrEP, living with HIV, STI diagnosis in the past 12 months, number of sex partners in the past 12 months, group sex in the past two months, chemsex in the past two months, and sex in sex venues or parties in the past two months. Except for living with HIV, all these potential covariables were also suggested as possible risk factors for mpox by ECDC [28]. A backward elimination procedure was employed in the multivariable regression models to retain only the significantly associated covariables. Covariable analyses were conducted on unweighted data, as potential covariables were used for data weighting. Odds ratios and adjusted odds ratios, along with their corresponding 95% CIs and p-values, were calculated for univariable and multivariable analyses. Additionally, Nagelkerke R-squared values were calculated as an approximation of the proportion of variation in the dependent variable that can be explained by the independent variables in a logistic regression model.

**Table 1:** Eligibility criteria for mpox (monkeypox) vaccination and related covariables included in the analyses.

| Eligibility criteria for vaccination against mpox amongst GBMSM |  |  |  | Covariables used in the analyses |
| --- | --- | --- | --- | --- |
| Eligibility criteria related to... | Netherlands mpox vaccination program 2022 |  | Derived from risk factors suggested by ECDC [28] |  |
|  | General eligibility criteria [23] | Specific eligibility criteria [22] |  |  |
| HIV-PrEP | People ‘*who use HIV-PrEP or are on the waiting list*’ | HIV-PrEP usage<br>PrEP waiting list | Use of HIV-PrEP<br>Eligibility for HIV-PrEP | Current use of HIV-PrEP<br>--- |
| HIV | People who ‘*are HIV-positive with a high risk of STIs*’ | Living with HIV<br>+ regular HCV screening | Not listed | Living with HIV<br>--- |
| STI-risk | People ‘*known to the sexual health centres (SHC) with a high risk of STIs*’ | Attending a sexual health centre (SHC) | Not listed | --- |
|  |  | + partner’s notification<br>or + having had an STI diagnosis | Recent history of bacterial sexually transmitted infections | ---<br>STI diagnosis (past 12 months) |
| Number of sex |  | or + >3 partners in the last six | Recent history of multiple sex | Number of male sex partners (past 12 |
| partners |  | months | partners or plans to engage with multiple partners | months) including 0–9, 10–19, and 20 or more. |
| Specific sexual activities |  | Not listed | Attending sex on premises venues<br>Group sex practices<br>Chemsex practices | Sex in sex venues and at parties (past two months)<br>Group sex (past two months)<br>Chemsex (past two months) |

The proportion of participants reporting one or more independent risk factors (IRFs) for mpox diagnosis, along with their 95% CIs, was calculated in the unweighted and weighted samples. Additional analyses were conducted to estimate the proportions of participants reporting specific numbers of IRFs, including one, two or three. The cumulative number of IRFs ranged from zero to three instead of four, as two of the four IRFs, namely current HIV-PrEP use and living with HIV, are mutually exclusive.

We estimated the proportion of individuals with immunity towards mpox due to previous mpox diagnosis, smallpox and mpox vaccination among participants with any independent risk factors for mpox. All participants diagnosed with mpox were considered to have achieved satisfactory immunity. Immunity through vaccination was also considered to be satisfactory if participants received two recent doses of the mpox vaccine. Note, however, that this was unlikely amongst participants as the survey was undertaken in the first month of the mpox vaccination program in the Netherlands. Immunity was also assumed to be satisfactory if participants received one dose of the mpox vaccine and had previously been vaccination through the historic smallpox vaccination program. Smallpox vaccination officially ended in November 1975 in the Netherlands (Manon Haverkate, Personal communication, 5 July 2023), and all participants born before 1976 were assumed to to have received smallpox vaccination in the past. Analyses related to immunity among participants were conducted on unweighted data only as the variables used for data weighting consisted of IRFs for mpox.

## Results

### Sample characteristics

Overall, 2,899 individuals accessed the survey, of whom 2,744 (94.7%) were adult GBMSM 18 years or over living in the Netherlands who provided informed consent to participate in the survey. Of the 2,744 eligible participants, 2,460 (89.7%) completed all questions and were included in the analyses. A comparison of sample characteristics before and after weighting is presented in Table 2. Sample age was slightly higher in the unweighted sample (Median = 42.0 years, IQR 23) than in the weighted sample (Median = 41.0, IQR 24). The weighting reduced the proportional representation of participants living in the Randstad metropolitan area (unweighted sample: 67.4%, weighted sample: 64.3%), and of the proportion of participants who self-identified as gay (unweighted sample: 91.5%, weighted sample: 88.9%). Reductions were also observed in the proportions of participants currently using HIV-PrEP (unweighted sample: 32.5%, weighted sample: 3.8%), living with HIV (unweighted sample: 11.0%, weighted sample: 4.3%), and diagnosed with an STI in the past 12 months (unweighted sample: 23.1%, weighted sample: 12.5%). The proportion of participants with 10–19 sex partners in the past 12 months was also reduced (unweighted sample: 18.4%, weighted sample: 15.0%), as was the proportion of participants with ≥20 sex partners in the past 12 months (unweighted sample: 23.9%, weighted sample: 14.3%). Reductions were also observed in the proportions of participants who, in the past two months, engaged in group sex (unweighted sample: 22.3%, weighted sample: 14.6%), chemsex (unweighted sample: 15.3%, weighted sample: 8.5%), or sex in sex venues or at parties (unweighted sample: 21.5%, weighted sample: 14.5%).

**Table 2:**
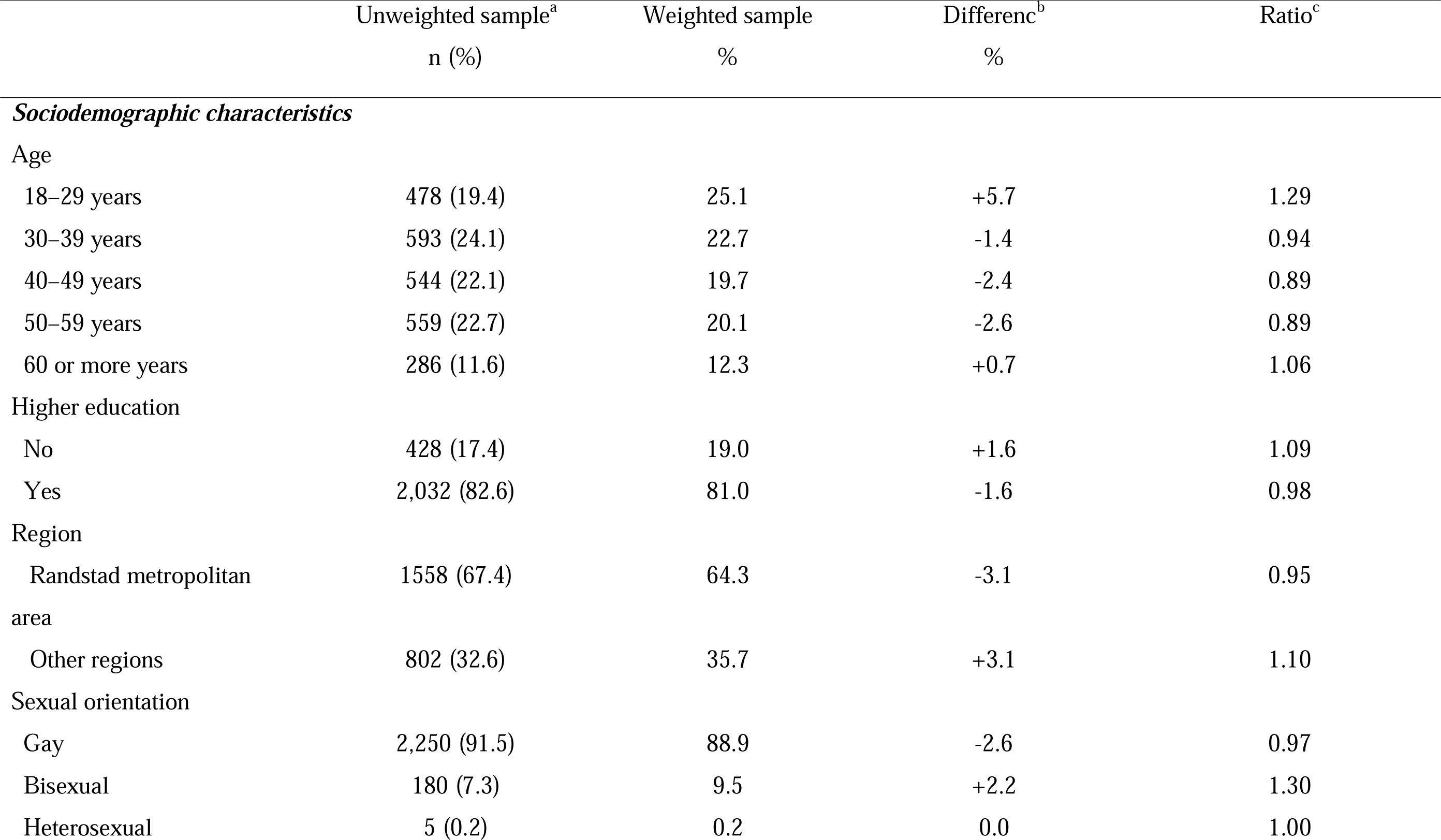
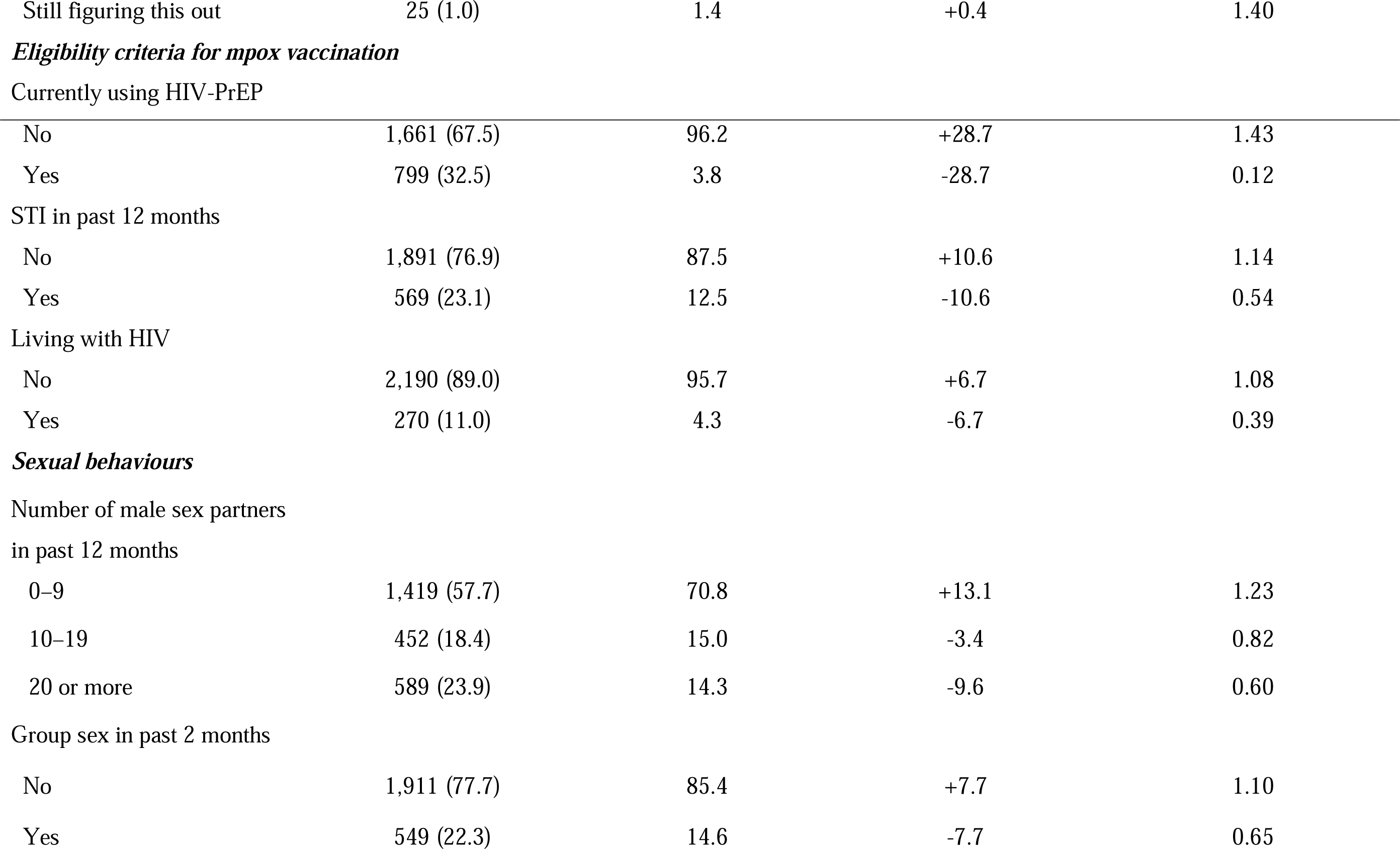
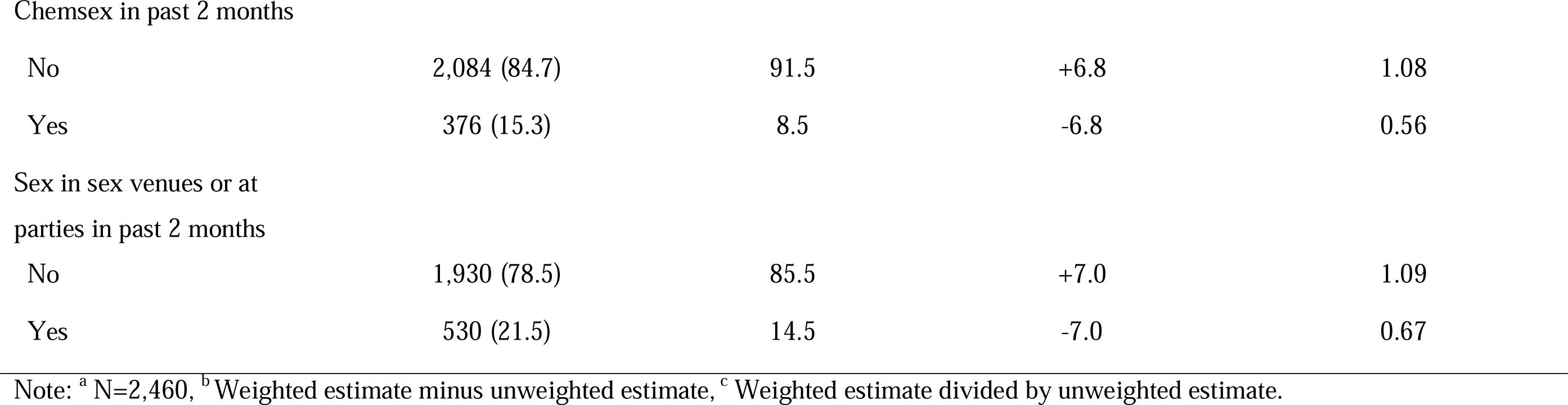
Participant characteristics – unweighted and weighted samples.

|  | Unweighted sample<sup>a</sup> | Weighted sample | Differenc<sup>b</sup> | Ratio<sup>c</sup> |
| --- | --- | --- | --- | --- |
|  | n (%) | % | % |  |
| *Sociodemographic characteristics* |  |  |  |  |
| Age |  |  |  |  |
| 18–29 years | 478 (19.4) | 25.1 | +5.7 | 1.29 |
| 30–39 years | 593 (24.1) | 22.7 | -1.4 | 0.94 |
| 40–49 years | 544 (22.1) | 19.7 | -2.4 | 0.89 |
| 50–59 years | 559 (22.7) | 20.1 | -2.6 | 0.89 |
| 60 or more years | 286 (11.6) | 12.3 | +0.7 | 1.06 |
| Higher education |  |  |  |  |
| No | 428 (17.4) | 19.0 | +1.6 | 1.09 |
| Yes | 2,032 (82.6) | 81.0 | -1.6 | 0.98 |
| Region |  |  |  |  |
| Randstad metropolitan area | 1558 (67.4) | 64.3 | -3.1 | 0.95 |
| Other regions | 802 (32.6) | 35.7 | +3.1 | 1.10 |
| Sexual orientation |  |  |  |  |
| Gay | 2,250 (91.5) | 88.9 | -2.6 | 0.97 |
| Bisexual | 180 (7.3) | 9.5 | +2.2 | 1.30 |
| Heterosexual | 5 (0.2) | 0.2 | 0.0 | 1.00 |
| Still figuring this out | 25 (1.0) | 1.4 | +0.4 | 1.40 |
| ***Eligibility criteria for mpox vaccination*** |  |  |  |  |
| Currently using HIV-PrEP |  |  |  |  |
| No | 1,661 (67.5) | 96.2 | +28.7 | 1.43 |
| Yes | 799 (32.5) | 3.8 | -28.7 | 0.12 |
| STI in past 12 months |  |  |  |  |
| No | 1,891 (76.9) | 87.5 | +10.6 | 1.14 |
| Yes | 569 (23.1) | 12.5 | -10.6 | 0.54 |
| Living with HIV |  |  |  |  |
| No | 2,190 (89.0) | 95.7 | +6.7 | 1.08 |
| Yes | 270 (11.0) | 4.3 | -6.7 | 0.39 |
| ***Sexual behaviours*** |  |  |  |  |
| Number of male sex partners<br>in past 12 months |  |  |  |  |
| 0–9 | 1,419 (57.7) | 70.8 | +13.1 | 1.23 |
| 10–19 | 452 (18.4) | 15.0 | -3.4 | 0.82 |
| 20 or more | 589 (23.9) | 14.3 | -9.6 | 0.60 |
| Group sex in past 2 months |  |  |  |  |
| No | 1,911 (77.7) | 85.4 | +7.7 | 1.10 |
| Yes | 549 (22.3) | 14.6 | -7.7 | 0.65 |
| Chemsex in past 2 months |  |  |  |  |
| No | 2,084 (84.7) | 91.5 | +6.8 | 1.08 |
| Yes | 376 (15.3) | 8.5 | -6.8 | 0.56 |
| Sex in sex venues or at parties in past 2 months |  |  |  |  |
| No | 1,930 (78.5) | 85.5 | +7.0 | 1.09 |
| Yes | 530 (21.5) | 14.5 | -7.0 | 0.67 |

Note: <sup>a</sup> N=2,460, <sup>b</sup> Weighted estimate minus unweighted estimate, <sup>c</sup> Weighted estimate divided by unweighted estimate.

### Rates and covariables of mpox diagnosis

Of the 2,460 participants in the unweighted sample, 73 (3.0%, 95% CI 2.3%–3.6%) reported that they had been diagnosed with mpox (see Table 3). The estimated proportion of mpox diagnoses in the weighted sample was 1.1% (95% CI 0.7% –1.6%).

**Table 3:** Rates of mpox diagnosis, invitation to vaccinate against mpox, and mpox vaccination.

|  | Unweighted sample <sup>a</sup> |  | Weighted sample | Difference<sup>b</sup> | Ratio<sup>c</sup> |
| --- | --- | --- | --- | --- | --- |
|  | n | % (95% CI) | % (95% CI) |  |  |
| Mpox diagnosis received | 73 | 3.0 (2.3–3.6) | 1.1 (0.7–1.6) | -1.9% | 0.37 |
| Invited for mpox vaccination | 708 | 28.8 (27.0–30.6) | 13.1 (11.8–14.4) | -15.7% | 0.45 |
| At least one dose of mpox vaccine received | 485 | 19.7 (18.1–21.3) | 7.8 (6.8–8.9) | -11.9% | 0.40 |

Note: <sup>a</sup> N=2;460; <sup>b</sup>Weighted percentage minus unweighted percentage; <sup>c</sup>Weighted percentage divided by unweighted percentage.

Results from regression analyses aimed at identifying the covariables of mpox diagnosis are presented in Table 4. In univariable analyses, mpox diagnosis was significantly associated with six potential covariables but not with living with HIV. All seven variables were included in the multivariate model, of which three were removed in the backward estimation (STI diagnosis in the past 12 months, group sex in the past two months and chemsex in the past two months) due to their lack of independent association with mpox diagnosis. In the final multivariable model, mpox diagnosis was significantly independently associated with current use of HIV-PrEP (aOR=3.92, 95% CI 2.01–7.66, p<.001) and living with HIV (aOR=2.67, 95% CI 1.15–6.22, p=.023). An independent association was also found with having had ≥20 sex partners in the past 12 months (aOR=5.16, 95% CI 2.24–10.68, p<.001) but with not having had 10-19 sex partners. Lastly, mpox diagnosis was independently associated with sex in sex venues or at sex parties in the past two months (aOR=2.11, 95% CI 1.26–3.52, p=.004). The multivariate model, however, only explained about a fifth of the variance in the risk of mpox diagnoses (Nagelkerke R-squared=0.18).

**Table 4:** Covariates of mpox diagnosis and mpox vaccination^a^.

|  | Recent mpox diagnosis |  |  |  |  | Recent mpox vaccination |  |  |  |  |
| --- | --- | --- | --- | --- | --- | --- | --- | --- | --- | --- |
|  | Rate | Univariable |  | Multivariable |  | Rate | Univariable |  | Multivariable |  |
|  |  | analyses |  | analysis |  |  | analyses |  | analysis |  |
|  | n (%) | OR | p-value | aOR | p-value | n/N (%) | OR | p-value | aOR | p-value |
|  |  | (95% CI) |  | (95% CI) |  |  | (95% CI) |  | (95% CI) |  |
| Current HIV-PrEP use |  |  |  |  |  |  |  |  |  |  |
| No | 22/1,661 (1.3) | Ref. |  | Ref. |  | 127/1,661 (7.6) | Ref. |  | Ref. |  |
| Yes | 51/799 (6.4) | 5.08 | <.001 | 3.92 | <.001 | 358/799 (44.8) | 9.81 | <.001 | 8.84 | <.001 |
|  |  | (3.06– |  | (2.01– |  |  | (7.80– |  | (6.71– |  |
|  |  | 8.44) |  | 7.66) |  |  | 12.32) |  | 11.66) |  |
| Living with HIV |  |  |  |  |  |  |  |  |  |  |
| No | 62/2,190 (2.8) | Ref. |  | Ref. |  | 439/2,190 (20.0) | Ref. |  | Ref. |  |
| Yes | 11/270 (4.1) | 1.46 | .259 | 2.67 | .023 | 46/270 (17.0) | .82 | .242 | 2.43 | <.001 |
|  |  | (.76– |  | (1.15– |  |  | (.59– |  | (1.63– |  |
|  |  | 2.80) |  | 6.22) |  |  | 1.14) |  | 3.60) |  |
| STI diagnosis<sup>b</sup> |  |  |  |  |  |  |  |  |  |  |
| No | 36/1,891 (1.9) | Ref. |  | --- |  | 260/1,891 (13.7) | Ref. |  |  |  |
| Yes | 37/569 (6.5) | 3.58 | p<.001 | --- | --- | 225/569 (39.5) | 4.10 | <.001 | 1.86 | <.001 |
|  |  | (2.24– |  |  |  |  | (3.32– |  | (1.45– |  |
|  |  | 5.73) |  |  |  |  | 5.08) |  | 2.39) |  |
| Number of sex partners<sup>b</sup> |  |  |  |  |  |  |  |  |  |  |
| 0-9 | 11/1419 (0.8) | Ref. |  | Ref. |  | 158/1419 (11.1) | Ref. |  | Ref. |  |
| 10-19 | 12/452 (2.7) | 3.49 |  | 1.98 | .118 | 124/542 (27.4) | 3.02 | <.001 | 1.54 | .005 |
|  |  | (1.53– |  | (.84– |  |  | (2.32– |  | (1.14– |  |
|  |  | 7.97) |  | 4.66) |  |  | 3.93) |  | 2.08) |  |
| 20 or more | 50/589 (8.5) | 11.87 | <.001 | 5.16 | <.001 | 203/589 (34.5) | 4.20 | <.001 | 1.65 | <.001 |
|  |  | (6.14– |  | (2.24– |  |  | (3.31– |  | (1.252.1 |  |
|  |  | 22.98) |  | 10.68) |  |  | 5.32) |  | 8) |  |
| Group sex<sup>c</sup> |  |  |  |  |  |  |  |  |  |  |
| No | 36/1,911 (1.9) |  |  | --- |  | 309/1,911 (16.2) |  |  | --- |  |
| Yes | 37/549 (6.7) | 3.76 | <.001 | --- | --- | 176/549 (32.1) | 2.45 | <.001 | --- | --- |
|  |  | (2.35– |  |  |  |  | (1.9– |  |  |  |
|  |  | 6.02) |  |  |  |  | 3.04) |  |  |  |
| Chemsex<sup>c</sup> |  |  |  |  |  |  |  |  |  |  |
| No | 46/2,084 (2.2) |  |  | --- |  | 357/2084 (17.1) |  |  | --- |  |
| Yes | 27/376 (7.2) | 3.43 | <.001 | --- | --- | 128/376 (34.0) | 2.50 | <.001 | --- | --- |
|  |  | (2.10– |  |  |  |  | (1.96– |  |  |  |
|  |  | 5.59) |  |  |  |  | 3.18) |  |  |  |
| Sex in venues or at parties<sup>c</sup> |  |  |  |  |  |  |  |  |  |  |
| No | 32/1,930 (1.7) |  |  |  |  | 329/1,930 (17.0) |  |  |  |  |
| Yes | 41/530 (7.7) | 4.97 | <.001 | 2.11 | .004 | 156/530 (29.4) | 2.03 | <.001 | --- | --- |
|  |  | (3.10– |  | (1.26– |  |  | (1.63– |  |  |  |
|  |  | 7.98) |  | 3.52) |  |  | 2.53) |  |  |  |

Note: <sup>a</sup> N=2,460; non-weighted sample only, <sup>b</sup> In the past 12 months, <sup>c</sup> In the past two months.

### Rates and covariables of mpox vaccination

Of the 2,460 participants in the unweighted sample, 708 (28.8%, 95% CI 27.0%–30.6%) had received an invitation to vaccinate against mpox, and 485 (19.7%, 95% CI 18.1%–21.3%) had received (initial) mpox vaccination (see Table 3). In the weighted sample, 13.1% (95% CI 11.8%–14.4%) had received an invitation for mpox vaccination, and 7.8% (95% CI 6.8%– 8.9%) had received (initial) mpox vaccination.

In univariable analyses, (initial) mpox vaccination was significantly associated with all potential covariables, except living with HIV (see Table 4). The backward procedure resulted in the elimination of three out of the seven terms from the model (group sex in the past two months, chemsex in the past two months and sex in sex venues or at sex parties in the past two months) due to their lack of independent association with mpox diagnosis. In the final multivariable model, mpox vaccination was significantly independently associated with current HIV-PrEP use (aOR=8.84, 95% CI 6.71–11.66, p<.001), living with HIV (aOR=2.43, 95% CI 1.63–3.60, p<.001), an STI diagnosis in the past 12 months (aOR=1.86, 95% CI 1.45–2.39, p<.001), having had 10-19 sex partners in the past 12 months (aOR=1.54, 95% CI 1.14–2.08, p=.005) and having had ≥20 sex partners in the past 12 months (aOR=1.65, 95% CI 1.25–2.18, p<.001). The multivariate model explained nearly a third of the variance in the uptake of mpox vaccination (Nagelkerke R-squared =.030).

### Proportion of the GBMSM subpopulation most at risk of mpox

The proportion of participants reporting zero, one, two or three IRFs for mpox diagnosis (i.e., current HIV-PrEP use or living with HIV, ≥20 sex partners in the past 12 months, or sex in sex venues or at parties in the past two months) is shown in Table 5.

**Table 5:**
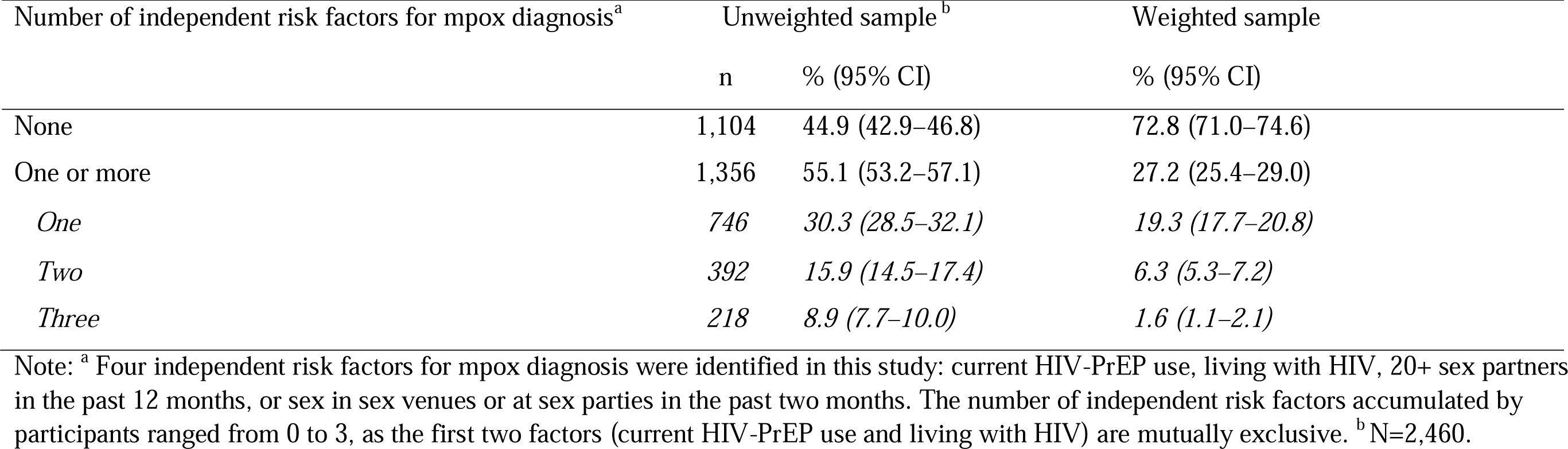
Proportions of GBMSM presenting independent risk factors for mpox diagnosis.

Of the 2,460 participants in the non-weighted sample, 1,104 (44.9%, 95% CI 42.9%–46.8%) presented no IRF, 746 (30.3%, 95% CI 28.5%–32.1%) presented one IRF, 392 (15.9%, 95% CI 14.5%–17.4%) two IRFs, and 218 (8.9%, 95% CI 7.7%–10.0%) three IRFs (Table 5). In the weighted sample, these proportions were 72.8% (95% CI 71.0%–74.6%), 19.3% (95% CI 17.7%–20.8%), 6.3% (95% CI 5.3%–7.2%), and 1.6% (95% CI 1.1%–2.1%), respectively. In total, 1,356 of the 2,460 participants in the non-weighted sample (55.1%, 95% CI 53.2%– 57.1%) presented at least one of the four IRFs for mpox diagnosis. This proportion was smaller (27.2%, 95% CI 25.4%–29.0%) in the weighted sample.

### Immunity among most-at-risk GBMSM

Of the 1,356 participants with one or more IRFs in the unweighted sample, 61 (4.5%, 95% CI 3.4%–5.6%) had been diagnosed with mpox, 181 (13.3%, 95% CI 11.5%–15.2%) had presumably received one dose of mpox vaccine while being vaccinated against smallpox, and 251 (18.5, 95% CI 16.4%–20.6%) had presumably received one dose of mpox vaccine but no smallpox vaccination (Table 6). Based on the proportions of participants reporting the first two situations, 17.8*%* (95% CI 15.8%–19.9%) of the 1,356 participants with one or more IRFs, would have presumably achieved a satisfactory immunity against mpox. The overall proportion of satisfactory immunity varied according to the number of independent risk factors reported, from 12.5% (95% CI 10.1%–14.8%) of the 746 participants with one IRF, 20.4% (95% CI 16.4%–24.4%) of the 392 participants with two IRFs and 31.7% (95% CI 25.4%–37.9%) of the 218 participants with three IRFs.

**Table 6:** Level of immunity towards mpox according to GBMSM’s numbers of independent risk factors for mpox diagnosis (unweighted data ^a^)

| Number of independent risk factors for mpox diagnosis<sup>a</sup> | n | Mpox diagnosis |  | Mpox vaccination <sup>b</sup> and smallpox vaccination <sup>c</sup> |  | Mpox vaccination <sup>b</sup> and no smallpox vaccination <sup>c</sup> |  | No mpox diagnosis nor mpox vaccination |  |
| --- | --- | --- | --- | --- | --- | --- | --- | --- | --- |
|  |  | n | % (95% CI) | n | % (95% CI) | n | % (95% CI) | n | % (95% CI) |
| None | 1,104 | 3 | 0.3 (0.0–0.6) | 12 | 1.1 (0.5–1.7) | 41 | 3.7 (2.6–4.8) | 1,048 | 94.9 (93.6–96.2) |
| One or more | 1,356 | 61 | 4.5 (3.4–5.6) | 181 | 13.3 (11.5–15.2) | 251 | 18.5 (16.4–20.6) | 863 | 63.6 (61.1–66.2) |
| *One* | 746 | 13 | 1.7 (0.8–2.7) | 80 | 10.7 (8.5–13.0) | 114 | 15.3 (12.7–17.9) | 539 | 72.3 (69.0–74.7) |
| *Two* | 392 | 18 | 4.6 (2.5–6.7) | 62 | 15.8 (12.2–19.4) | 84 | 21.4 (17.4–25.4) | 228 | 58.2 (53.3–63.1) |
| *Three* | 218 | 30 | 13.8 (9.2–18.4) | 39 | 17.9 (12.8–23.0) | 53 | 24.3 (18.6–30.1) | 96 | 44.0 (34.7–50.7) |

Note: <sup>a</sup> N=2,460. <sup>b</sup> Due to the timing of the survey, we assumed that all GBMSM reporting mpox vaccination had only received one dose of vaccine. <sup>c</sup> All participants born before 1976 were assumed to have received smallpox vaccination.

## Discussion

Our findings provide novel insights into the extent and covariables of the risk of mpox diagnosis as well as the uptake and need of mpox vaccination among GBMSM in the Netherlands. Of the participants in this study, 3.0% had been diagnosed with mpox and a fifth (19.7%) had received (initial) mpox vaccination since the start of the mpox outbreak. Using data weighting to attenuate participation bias, we found that an estimated 1.1% of the sexually active GBMSM population 18-80 years in the Netherlands had been diagnosed with mpox and less than a tenth (7.8%) had received (initial) mpox vaccination. Our estimates seem to surpass those derived from the national records of mpox cases and first dose mpox vaccination at the time the survey recruitment ended. By September 1st, 2022, 1,080 GBMSM mpox cases [11] and 12,820 first vaccine doses [29] had been recorded nationally. Extrapolating these figures to a population of 310,000 GBMSM yields a mpox prevalence estimate of 0.3% and a vaccination prevalence estimate of 4.1%.

Our multivariable assessment of covariables found commonalities and differences in the independent covariables of mpox diagnosis and mpox vaccination. Mpox diagnosis and mpox vaccination were each independently associated with HIV-PrEP use, living with HIV and reporting ≥20 sex partners in the past 12 months. STI diagnosis in the past 12 months and 10– 19 partners in the past 12 months were independently associated with mpox vaccination but not with mpox diagnosis. Sex in sex venues or at parties in the past two months was independently associated with mpox diagnosis but not with mpox vaccination. Neither group sex nor chemsex in the past two months were independently associated with mpox diagnosis or vaccination, although associations were observed in univariable analyses.

In settings with limited mpox vaccine availability, mpox vaccination is best targeted at GBMSM most at risk for mpox, as also recommended by ECDC [28], and our findings provide novel guidance to further improve the alignment between the eligibility criteria for mpox vaccination and the evidence regarding risk factors for mpox. Our findings underscore the importance of four IRFs for mpox diagnosis: current use of HIV-PrEP, living with HIV, a higher number (≥20) of sex partners in the past 12 months, and having sex in sex venues or at sex parties in the past 2 months.

These findings confirm the importance of HIV-PrEP use as an eligibility criterion for mpox vaccination among GBMSM in the Netherlands [22], and more broadly in Europe [28]. Our results also suggest the importance of maintaining living with HIV as eligibility criterion in the Netherlands [22], although this was not listed by ECDC [28]. The findings also provide new insights into the ways in which sexual activity affects GBMSM’s vulnerability to mpox. The finding that both a high number of sex partners and having sex in sex venues or parties were independently associated with mpox diagnosis is noteworthy. This suggests that GBMSM’s susceptibility to mpox does not merely result from their number of sex partners (and sex contacts) but also from engaging in specific sexual activities and networks.

The findings on IRF for mpox diagnosis can help refine the eligibility criteria for mpox vaccination, starting with the criteria used in the Netherlands. Having multiple sex partners was noted as a potential risk factor for mpox in ECDC guidance [28]. During the 2022 vaccination campaign in the Netherlands, the number of partners was not used as a primary eligibility criteria. However, GBMSM known from the sexual health centres as being at higher risk of STIs had to have more than 3 partners in the last 6 months to be invited for vaccination [22]. In light of our results on the number of partners from which the risk for mpox become significantly higher, the threshold of three partners per semester seems relatively low. In the context of limited vaccine stock, we suggest using ≥10 partners in the past 6 months as an independent eligibility criterion for vaccinating most at risk GBMSM in the Netherlands. This criterion derives from an adaptation of the ≥20 partners threshold established in this study to the 6-month timeframe used for the recording national indicator data.

Our findings also confirm ECDC guidance that attending sex venues is a risk factor for mpox [28], and this should be included as an eligibility criterion for mpox vaccination among GBMSM in the Netherlands. However, unlike suggested by ECDC [28], we did not find that engaging in group sex, chemsex, or having a recent history of STI were independent risk factors for mpox. Nevertheless, as these variables were associated with mpox diagnosis in univariate analyses, they could potentially offer additional insights in determining the need for mpox vaccination, particularly when data concerning other eligibility criteria, such as having sex at sex venues or parties, are not available.

Our assessment enabled estimating the proportions of GBMSM most at risk for mpox and in need of vaccination. In the non-weighted sample, more than half of the participants (55.1%) presented at least one of the four IRFs for mpox. This proportion was reduced to over a quarter (27.2%) in the weighted sample, equating to approximately 83,700 individuals when extrapolated to an estimated 310,000 adult GBMSM.

As the number of IRFs accumulated increased, the size of the high-risk groups decreased. Specifically, the presumed core transmission group, consisting in this study of GBMSM with three IRFs, accounted for less than a tenth (8.9%) of participants in the non-weighted sample and less than 2% (1.6%) in the weighted sample. The study also contributed to estimating the proportion of GBMSM at higher risk who had achieved a satisfactory level of immunity towards mpox at the time of the survey. Among the participants with one or more IRFs, less than one in five (17.8%) had satisfactory immunity either due to infection or vaccination. The proportion of satisfactory immunity varied according to the number of IRFs reported, from above a tenth (12.5%) among participants with one IRF, to a fifth (20.4%) among participants with two IRFs and near a third (31.7%) among participants with three IRFs. If all GBMSM born after 1975, who have not been vaccinated against smallpox but had presumably received only one dose of mpox vaccine were to receive a second dose — acknowledging the challenges of achieving full compliance with the recommendation to obtaining two doses — the resultant immunity levels would approximate 27.8%, 41.8%, and 56% among GBMSM with one, two, and three IRFs for mpox, respectively.

These findings show that the level of immunity against mpox achieved at the time of the survey and potentially achieved if second doses are received after the time of the survey, was highest amongst GBMSM reporting three IRFs. As modelling has shown [15], a vaccination strategy targeting those most at risk may contribute to effectively curbing mpox outbreaks. It is, however, possible that mpox is reintroduced among lower risk and less vaccinated and immune GBMSM, for instance those who present one or two IRFs for mpox. Vaccination of at least all GBMSM at risk presenting one or more IRFs is hence needed [21].

This study has unique strengths. To the best of our knowledge, this is the first study to assess the extent of mpox diagnoses and vaccination in a community sample of GBMSM, and to assess and compare commonalities and differences in covariables for mpox diagnosis and mpox vaccination. Also, we recruited a large sample of GBMSM and achieved a high survey completion rate. Furthermore, we provided an updated estimate of the size of the sexually active adult GBMSM population in the Netherlands and used weighting procedures to reduce some recruitment biases. Our study also has limitations that are common to survey research in GBMSM, including potential memory and social desirability biases related to self-report. We mitigated these biases by clearly specifying reporting periods and ensuring anonymity of responses. Furthermore, we used convenience sampling and, despite the use of weighting procedures, the findings may not be representative of the population of GBMSM in the Netherlands. The survey’s focus may have led to an overrepresentation of GBMSM interested in mpox, including individuals who had mpox or had obtained vaccination. Furthermore, as mpox is a relatively rare condition, the number of participants diagnosed with mpox was limited, despite the large sample size. This low number led to the risk of not being able to detect associations with covariables of mpox diagnosis. Our analyses on covariables of mpox diagnosis would therefore need to be replicated in a sample that includes a larger number of participants diagnosed with mpox, including people living with HIV.

Also, some information on specific eligibility criteria for mpox vaccination was not assessed in the survey, notably being on a waiting list for PrEP and regular HCV screening among participants living with HIV. In addition, the survey did not allow for differentiating between vaccination pre- or post-exposure to mpox, albeit that uptake of post-exposure vaccinations remains very limited in the Netherlands. Furthermore, the survey assessed mpox diagnoses and vaccination uptake and covariables only during August 2022, which was merely one month into the vaccination program. However, while the reach of the vaccination program has increased thereafter, vaccinations mostly occurred at the start of the program, and GBMSM at highest risk for mpox were invited first.

The prospect of mpox reintroduction from endemic regions remains a possibility, carrying with it the potential for novel mpox outbreaks. To prevent future reintroductions from giving rise to new outbreaks, and to ensure the sustained eradication of mpox [18], even in the absence of such introductions, the vaccination of all GBMSM, and any other groups at risk for mpox, may prove necessary. However, the comprehensive vaccination of entire groups or populations can be constrained by factors such as limited vaccine availability, lack of (human) resources, or a little enthusiasm for mpox vaccination among the targeted groups or population. In situations where universal vaccination is not feasible, our research findings play a crucial role in determining individuals most susceptible to mpox, thereby offering a targeted approach to mpox vaccination.

The findings of our study underscore the importance of an evidence-based approach to identifying risk factors for mpox diagnosis and guiding the selection of eligibility criteria for mpox vaccination. Our approach has already contributed to a refinement of vaccination eligibility criteria applied as of May 2023 in the Netherlands [16]. Both ≥10 partners in the past six months and having sex at sex venues or parties were added to the list of eligibility criteria [16]. Our findings have also contributed to the policy change to provide access to vaccination to a larger number of GBMSM than initially planned [30]. As of May 2023, unvaccinated GBMSM who self-identify as high-risk for mpox and GBMSM born after 1975 who received only a single vaccine dose have the possibility to contact their healthcare service to discuss mpox vaccination[16]. This extension of the scope of the vaccination program will provide valuable health benefits to a larger group of GBMSM at risk of mpox.

## Data availability

Data are available from the corresponding author upon reasonable request.

## Acknowledgements

The authors thank Manon Haverkate, Lisette Kuyper, and Hanneke de Graaf for sharing information and insights, Arjan van Bijnen and Laurian Kuipers for their contribution to the recruitment, and all participants to the survey.

## Authors’ contributions

PA developed the idea for this study, conducted the statistical analyses, and wrote the first draft of the manuscript together with JdW. EOdC and MX provided important additions to the manuscript, including information and insights on the mpox outbreak and vaccination program in the Netherlands. PZ, HB, CB, IK, SD, and MtW reviewed the manuscript and contributed further revisions. All authors have read the final manuscript and accepted authorship.

