## Supplementary material for "An assessment of rates and covariates of mpox diagnosis and vaccination provides evidence to refine eligibility criteria for mpox vaccination among gay, bisexual and other men who have sex with men in the Netherlands"

Adam PCG, ([https://orcid.org/0000-0002-8979-7372](https://orcid.org/0000-0002-8979-7372?lang=en)), Op de Coul ELM (<https://orcid.org/0000-0002-8263-9187>), Zantkuijl P (https://orcid.org/ 0000000328388767), Xiridou M (<https://orcid.org/0000-0002-5257-4503>), Bos H, Blom C, Ketsuwan I, te Wierik MJM, David S, de Wit JBF (<https://orcid.org/0000-0002-5895-7935>).

**Supplementary material**

This supplementary material details the data and assumptions used to provide an updated and adjusted estimate of the size of the currently sexually active GBMSM population 18-80 years in the Netherlands in 2022.

**Estimating the size of the adult GBMSM population in the Netherlands**

Our analyses required a baseline estimate of the number of sexually active adult GBMSM living in the Netherlands. The main source of information on the size of the GBMSM population in the Netherlands is an assessment derived from a 2006 population-based survey that collected data on the proportions of adult males who were attracted to men, ever had sex with them, or self-identified as gay or bisexual [1]. According to this prior estimate, 16% of males aged 19-69 in the Netherlands in 2006 reported affirmatively on at least one of the three aspects of gay or bisexual experience, and 6% of all males aged 19-69 reported affirmatively on all three aspects. Extrapolating the 6% prevalence estimate to the size of the male population 19-69 years old living in the Netherlands in 2022 (N= 5,805,000) gives an estimated 348,300 males aged 19-69 years who are attracted to men, ever had sex with men and self-identified as gay or bisexual in that year. To derive an estimate of the size of the population of interest for our study (i.e., sexually active GBMSM aged 18-80 years in the Netherlands), additional adaptations relating to age and sexual activity are, however, required.

**Correcting the adult GBMSM population estimate for missing age groups**

At the time we finalised the manuscript, GBMSM diagnosed with MPXV infection in the Netherlands were aged 18 to 77 years old [2], which is close to the age range of participants in our study (18-80 years). As the previously estimated 6% prevalence of men who are attracted to men, ever had sex with men and self-identified as gay or bisexual pertained to individuals aged 19-69 years only [1], we further estimated the proportion of individuals reporting affirmatively on all three aspects of gay or bisexual experience among males aged 18 and 70-80 years of age, respectively.

*Males aged 18 years old:* According to a 2017 population survey, half of young people aged 12-25 years in the Netherlands have had sexual intercourse by the age of 18.6 years [3,4]. We assumed there was no difference in age of sexual debut according to sexual orientation. As not all young men start having sex before the age of 19, we assumed that the rate of men 18 years of age reporting gay or bisexual attraction, behaviour and identity could be lower than the 6% estimated for the 19-69 years old [1]. We, therefore, set the rate estimate of males aged 18 years who would respond affirmatively on all three aspects of gay or bisexual experience to 4.5% (see Table 1).

*Males aged 70-80 years old:* The comparatively high rate of same-sex experiences in the Netherlands is attributed to the tolerant attitudes towards homosexuality that has existed for several decades [5]. A less tolerant social environment may have existed in the past, which may have prevented some individuals in older generations from acknowledging and/or enacting same-sex attraction, behaviour or identity. We therefore reduced the estimated rate of males aged 70-80 years who present all three aspects of gay or bisexual experience to 5% (see Table 1).

**Correcting the estimated GBMSM population size for recent sexual activity**

The previous estimate of the size of the GBMSM population in the Netherlands encompassed ever having had sex with men as criterion for inclusion in the GBMSM population, in addition to same-sex attraction and self-identified as gay or bisexual [1]. The resulting estimate of the size of the GBMSM population therefore includes individuals who have not been recently sexually active with men. These individuals are not the focus of our study as they are not at risk of MPXV infection, nor are they in need of mpox vaccination. We, therefore, made additional corrections to restrict the estimated size of the GBMSM population to those who have recently been sexually active (i.e. who engaged in sexual activity at least once in the past six months). For this estimation, we assumed that, among males reporting same-sex attraction, ever having had sex with a man and self-identification as gay or bisexual, the proportion who recently were sexually active would be 75% for males aged 18 years, 85% for males 19-54 years old, 75% for males 55-69 years old, 60% for males 70-74 old, and 50% for males 75-79 years old (see Table 1). Using these proportions, we estimated for each age group the proportion of GBMSM 18-80 years who had been recently sexually active, and subsequently estimated the proportion of males who are attracted to men, recently had sex with men, and self-identified as gay or bisexual to be 3.4% for males 18 years, 5.1% for males 19-54 years, 4.5% for males 55-69 years, 3.0% for males 70-74 years, and 2.5% for males 75-79 years.

Table S1: Estimates of the 2022 proportion of sexually active men reporting same-sex attraction, sex with men and gay or bisexual identity, by age group.

| Age group | Estimated proportion of men reporting same sex attraction, ever sex with men, and gay or bisexual identity | Proportion of GBMSM reporting being recently sexually active | Estimated proportion of men reporting same sex attraction, recent sex with men, and gay or bisexual |
| --- | --- | --- | --- |
| 18 | 4.5% | 75% | 3.4% |
| 19-34 | 6% [1] | 85% | 5.1% |
| 35-54 | 6% [1] | 85% | 5.1% |
| 55-69 | 6% [1] | 75% | 4.5% |
| 70-74 | 5% | 60% | 3.0% |
| 75-80 | 5% | 50% | 2.5% |

**Size of the sexually active adult GBMSM population in the Netherlands**

By extrapolating the corrected estimates for the proportion of sexually active adult GBMSM by age group to the estimated size of each of these age groups within the male population 18-80 years old in the Netherlands in 2022 (see Table 2), we estimated that there would be 312,918 males who were attracted to men, recently had sex with men, and self-identified as gay or bisexual out of a total population of 6,760,000 males aged 18-80 years old in the Netherlands.

Table S2: Size of the sexually active GBMSM population per age group.

| Age group | Number of males  in the general population [6] | Recently sexually active GBMSM | |
| --- | --- | --- | --- |
|  |  | Proportion | Number |
| 18 | 108,000 | 3.40% | 3672 |
| 19-34 | 1,845,000 | 5.10% | 94095 |
| 35-54 | 2,246,000 | 5.10% | 114546 |
| 55-69 | 1,714,000 | 4.50% | 77130 |
| 70-74 | 460,000 | 3.00% | 13800 |
| 75-80 | 387,000 | 2.50% | 9675 |
| Total | 6,760,000 | 4.63% | 312918 |

4.	Rutgers [Internet]. Utrecht, the Netherlands: Rutgers. Young people in the Netherlands start having sex at a later age; 2017 Jun 17 [cited 2023 Feb 24]; [about 5 screens].

<https://rutgers.international/news/young-people-in-the-netherlands-start-having-sex-at-a-later-age/> (Retrieved 21/12/2022)

5.	Kuyper L, Vanwesenbeeck I. High levels of same-sex experiences in the Netherlands: Prevalences of same-sex experiences in historical and international perspective. J Homosex. 2009;56(8):993–1010. <https://doi.org/10.1080/00918360903275401>

6.	Statline [Internet]. Den Haag, the Netherlands: Statistics Netherlands c2023. Bevolking; kerncijfers [Population; key numbers]; 2022 Aug 22 [cited 2023 Feb 26]; [about 1 screen]. Available from: <https://opendata.cbs.nl/statline/#/CBS/nl/dataset/37296ned/table?ts=1677400975218>
